# Type A Aortic Dissection Repair at High Versus Low Volume Centers

**DOI:** 10.1101/2025.05.02.25326910

**Authors:** Aryan Meknat, Alee N. Pettit, Peter Cho, Neusha Hollingsworth, Whitney Taylor, Michael S. Halbreiner

## Abstract

**Background:** Studies have shown significant improvement in outcomes when ATAAD repairs are performed at high-volume centers, with both center and surgeon volumes contributing to improved survival. The aim of our study was to evaluate our own network and determine differences in ATAAD outcomes between our high- and low-volume aortic centers.

**Methods:** This was an observational, multi-center retrospective study consisting of 205 cases of ATAAD repair within our institution, consisting of 3 hospitals within the region that perform cardiac surgery, from January 2017 to January 2025. Preoperative characteristics, operative characteristics, and postoperative outcomes were collected, stratified by center volume (high vs low) and analyzed.

**Results:** There were 205 patients identified with ATAAD who presented to our health network. Of these, 164 presented to our high-volume center while 41 presented to our low volume centers. When stratified by center volume, there was no significant difference in preoperative characteristics. What was significant were the cardiopulmonary bypass (CPB) [174 (137-218) versus 236.5 (195.5-288) minutes, p <0.001], circulatory arrest [30 (22-45) versus 45 (33-67) minutes, p=0.001], and cross clamp times [93 (72-127) versus 131 (94.5-194) minutes, p=0.002]. The univariable, survival analysis did show a significant difference in survival at 3 years – 81.5% versus 66.7% [p=0.009]. Utilizing a multivariable Cox regression model, having surgery at a high-volume center was found to be associated with a significant difference in 3-year survival [p=0.021].

**Conclusion:** Time from presentation to surgery influences mortality, but overall mortality has been shown to be much more dependent on where the repair is performed.

**Central Message:** Studies, including our own, have shown significant improvement in outcomes when dissection repair is performed at high-volume centers because of technical innovations and multi-disciplinary expertise.

**Perspective Statement:** A regional care model with emphasis on diagnosis and treatment protocols has been shown to reduce times to diagnosis and treatment, with both center and surgeon volumes contributing to improved aortic dissection survival. The aim of our study was to evaluate our own network and determine differences in acute type A aortic dissection outcomes between our high- and low-volume aortic centers.

## Background

Despite advances in medical and surgical management of aortic disease, acute type A aortic dissections (ATAAD) continue to be a life-threatening condition with an incidence of 2-16 cases per 100,000 patient years. Up to 50% of patients die before reaching a hospital, making immediate surgical repair still the standard of care.^1^ The mortality rate of ATAAD during the initial 24 to 48 hours after symptom onset is still discussed and described as 1% to 2% per hour, which was based on old dogma. ATAAD data from the 1950s estimated mortality at 21% in the first 24 hours and 37% in the first 48 hours (1% – 2% per hour), before medical therapy improved, and cardiovascular surgery became routine.^2^ In a more recent study using International Registry of Acute Aortic Dissection data, nonoperative, medically managed ATAADs had a 48-hour mortality of 23.7%, or 0.5% per hour, while surgically managed ATAADs led to a mortality of 4.4%, or 0.09% per hour.^3^ Another study found an hourly mortality rate during the first 24 hours to be 2.6% in nonsurgical ATAAD.^1^ These studies emphasize the importance of quick diagnosis, transfer, and treatment of type A aortic dissections.

Several studies have shown significant improvement in outcomes when ATAAD repairs are performed at high-volume centers because of technical innovations and multi-disciplinary expertise. One study showed in-hospital mortality of 14.0% if a high-volume aortic surgeon was present versus a 24.0% in-hospital mortality when a low-volume aortic surgeon present. The mortality odds ratio in this study for a low-volume aortic team was 3.72 (p = .01).^4^ Another study showed a significant reduction in operative mortality when low-volume hospitals were compared with high-volume hospitals (27% vs 16%; *P* < .001).^5^ A regional care model with emphasis on diagnosis and treatment protocols has been shown to reduce times to diagnosis and treatment, with both center and surgeon volumes contributing to improved aortic dissection survival. Therefore, rapid transfer to aortic centers with the appropriate multi-disciplinary team adept at complex aortic surgery, cerebral protection strategies, and techniques to address malperfusion is critical to optimize outcomes.

Interhospital transfer is needed in more than 70% of cases, leading to inherent treatment delays. But in a study where a 24/7 dissection ‘hotline’ was introduced for referring centers to transfer to 3 centers with experienced aortic teams, in-hospital mortality significantly reduced from 24.2% to 11.11% and 30-day mortality was significantly reduced from 20% to 8%.^6^ Another study showed that despite delaying surgery, transferring patients to high-volume hospitals was associated with 7.2% absolute risk reduction in operative mortality.^7^ Furthermore, according to the 2022 ACA/AHA Aortic Guidelines, despite an inherent delay in the start time of surgery, transfer from low- to high-volume hospitals (one that performs ≥7 aortic root, ascending aortic, or aortic dissection repairs per year), as part of regionalization of care, can result in significantly improved outcomes. The 2021 AATS expert consensus document on ATAADs suggests that ATAAD complicated by comorbidities, previous cardiac surgery, or distal malperfusion are candidates for escalation of care to a comprehensive aortic center (CAC). Also, certain surgical and endovascular therapies require an additional level of expertise that might warrant transfer to a CAC. The effect of case volumes has consistently shown that higher-volume centers achieve better patient survival after ATAAD repair compared with lower-volume centers. The aim of our study was to evaluate our own network and determine differences in ATAAD outcomes between our high- and low-volume aortic centers.

## Methods

This was an observational, multi-center retrospective study consisting of 205 cases of ATAAD repair within our institution, consisting of 3 hospitals within the region that perform cardiac surgery, from January 2017 to January 2025. We obtained Institutional Review Board approval for this study. Preoperative characteristics, operative characteristics, and postoperative outcomes were collected, stratified by center volume (high vs low) and analyzed. There is a single high-volume center (>1000 cardiac surgery cases per year and >130 aortic surgeries), while our low volume centers included two other hospitals within our network that do not routinely perform aortic procedures (i.e. less than 10 cases year) but that do perform ATAAD repairs. Data was presented as median (interquartile range) for continuous measures and n (%) for categorical measures. There were p-values for Wilcoxon rank-sum test or Pearson’s chi-squared tests for continuous and categorical variables, respectively. Statistical significance was defined by p ≤ 0.05.

Three-year survival post-operation was compared between high and low volume groups. The Kaplan-Meier method was used to construct 3-year survival curves, which was stratified by volume, with 95% confidence interval displayed in Figure 2. The significance of survival difference was calculated using the log-rank test. We also constructed a multivariable Cox regression model – to identify independent predictors of 3-year survival decrement. Forward stepwise selection (p<0.20) was used to select variables from pre-operative and operative characteristics. For survival analyses, patient data were censored at the time of 3-year follow-up. The pre-operative characteristic data was modeled after the German registry for acute aortic dissection type A score (GERAADA) – to determine dissection severity presenting to our high versus low centers.^8, 9^ Operative characteristics were focused on factors that could affect post-operative complication rates and overall outcomes – type of repair(s) (e.g. hemiarch vs arch +/- root replacement), cannulation strategy, and cardiopulmonary bypass; cross-clamp; and circulatory arrest times. Postoperative outcomes data included a myriad of complications that could arise after any cardiac surgery but also could be a product of malperfusion arising from ATAAD. Another point of emphasis was intensive care unit (ICU) and overall hospital length of stay, as well as mortality rate – 30-day, 90-day, and 3-year survival. All analyses in this study were performed using STATA 18.0 (StataCorp, College Station, TX).

**Figure 1.**
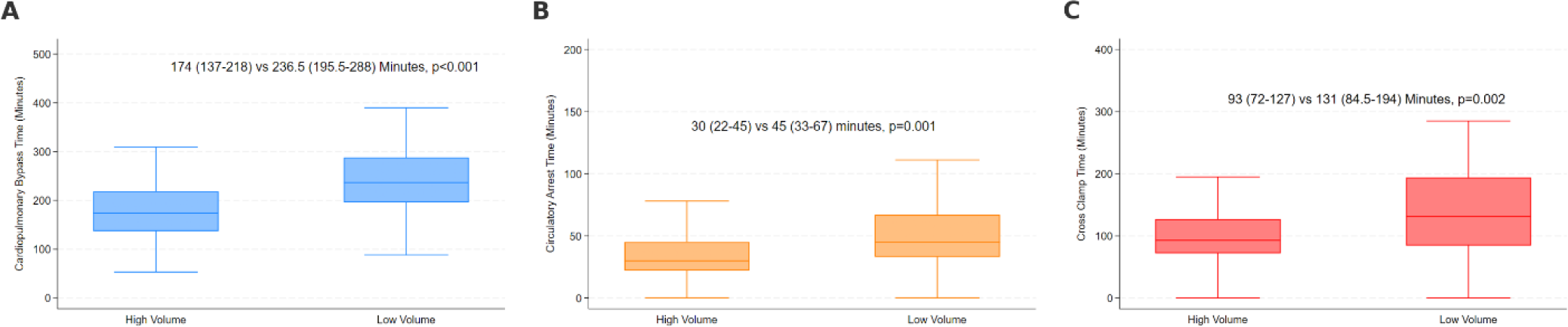
A: Cardiopulmonary Bypass Time, B: Circulatory Arrest Time, C = Cross Clamp Time.

**Figure 2.**
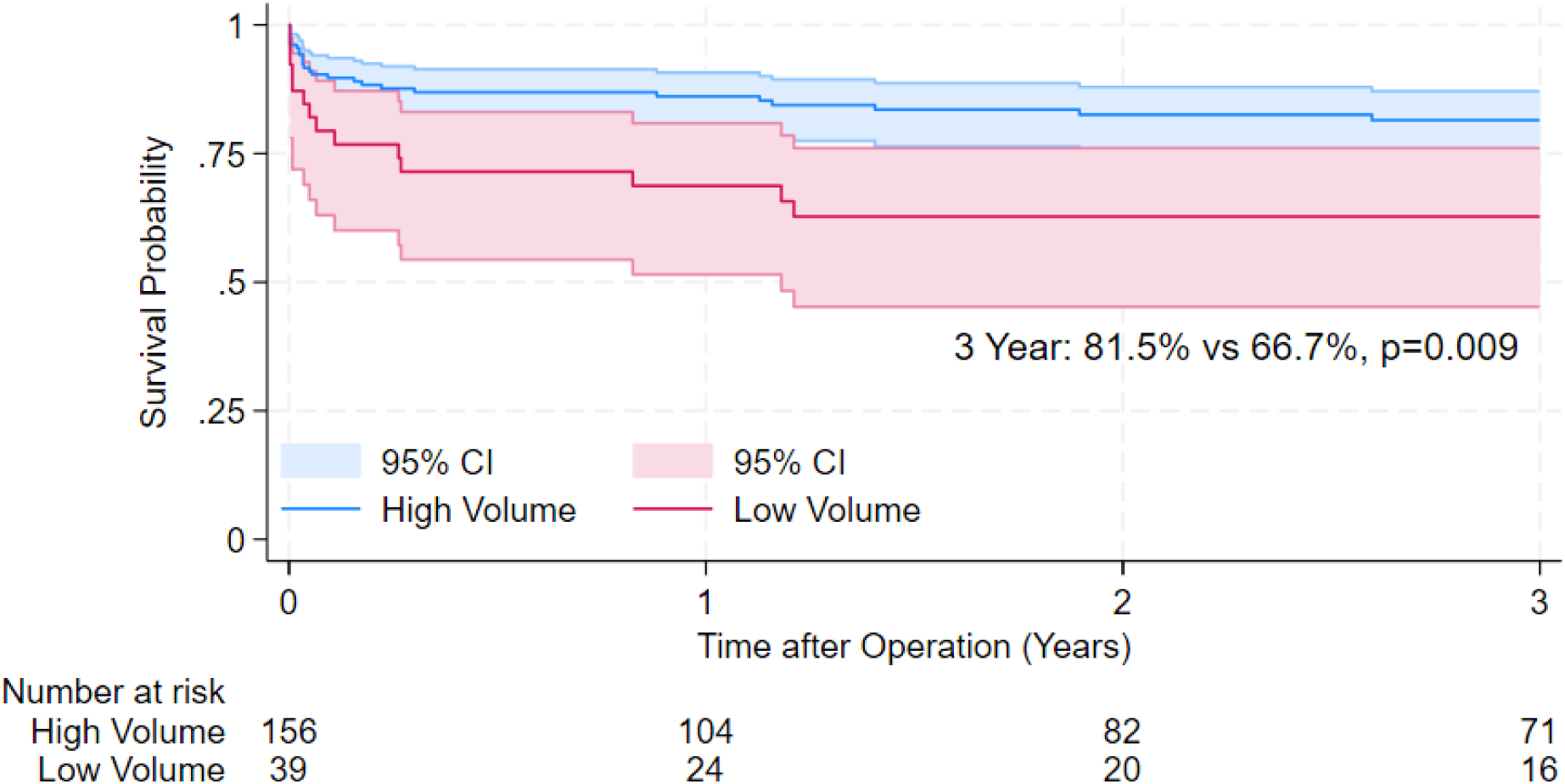
Kaplan-Meier Curve Stratified by Center Volume for Aortic Dissection Repair

## Results

There were 205 patients identified with ATAAD who presented to our health network. Of these, 164 (80%) presented to our high-volume center while 41 (20%) presented to our low volume centers. When stratified by center volume, there was no statistically significant difference in preoperative characteristics (Table 1). Despite not being statistically significant there were a greater number of patients who presented with cerebral [11 [6.7%] versus 1 [2.4%], p=0.3] and spinal cord malperfusion [8 [4.9%] versus 0 [0%], p=0.15], at the high-volume center. Also noteworthy were the number of patients that presented with inotropic and/or vasopressor support at the high versus low volume centers, 23 [14.2%] versus 2 [4.9%], respectively [p=0.1].

**TABLE 1.** Pre-operative Characteristics Stratified by Center Volume.

|  | **High**<br>**N=164** | **Low**<br>**N=41** | **p-value** |
| --- | --- | --- | --- |
| Age (years) | 67 (57.5-74.5) | 67 (54-73) | 0.68 |
| Gender |  |  | 0.44 |
| Male | 99 (60.4%) | 29 (70.7%) |  |
| Female | 64 (39.0%) | 12 (29.3%) |  |
| Race |  |  | 0.60 |
| White | 134 (81.7%) | 34 (82.9%) |  |
| Black | 22 (13.4%) | 4 (9.8%) |  |
| Asian | 2 (1.2%) | 0 (0.0%) |  |
| Other | 6 (3.7%) | 3 (7.3%) |  |
| Previous Cardiac Surgery | 23 (14.1%) | 8 (19.5%) | 0.39 |
| Previous Sternotomy | 19 (11.6%) | 6 (14.6%) | 0.59 |
| Aortic Aneurysm | 29 (17.7%) | 8 (19.5%) | 0.78 |
| Organ Malperfusion | 49 (29.9%) | 14 (34.1%) | 0.60 |
| Coronary | 7 (4.3%) | 3 (7.3%) | 0.42 |
| Cerebral | 11 (6.7%) | 1 (2.4%) | 0.30 |
| Spinal cord | 8 (4.9%) | 0 (0.0%) | 0.15 |
| Mesenteric | 7 (4.3%) | 0 (0.0%) | 0.18 |
| Renal | 15 (9.1%) | 7 (17.1%) | 0.14 |
| Hepatic | 3 (1.8%) | 0 (0.0%) | 0.38 |
| Limb (Vascular) | 22 (13.4%) | 6 (14.6%) | 0.84 |
| Inotrope and Vasopressor | 23 (14.2%) | 2 (4.9%) | 0.10 |
| Beta Blocker | 75 (45.7%) | 22 (53.7%) | 0.36 |
| Calcium Channel Blocker | 33 (20.1%) | 5 (12.2%) | 0.24 |
| Mechanical Intubation | 14 (8.6%) | 1 (2.4%) | 0.18 |
| Required Cardiopulmonary Resuscitation | 5 (3.0%) | 0 (0.0%) | 0.26 |
| Lactate (mmol/L) | 1.6 (1-2.7) | 2.4 (1.2-4.3) | 0.10 |
| Left Ventricular Ejection Fraction | 60 (55-65) | 60 (60-60) | 0.59 |
| Aortic Valve Insufficiency | 36 (57%) | 9 (64%) | 0.81 |
| Mild | 23 (64%) | 7 (78%) |  |
| Moderate | 7 (19%) | 2 (22%) |  |
| Severe | 5 (14%) | 0 (0%) |  |

Data are presented as median (IQR) for continuous measures, and n (%) for categorical measures. P-values for Wilcoxon rank-sum test or Pearson's chi-squared tests for continuous and categorical variables, respectively.

**TABLE 2.**
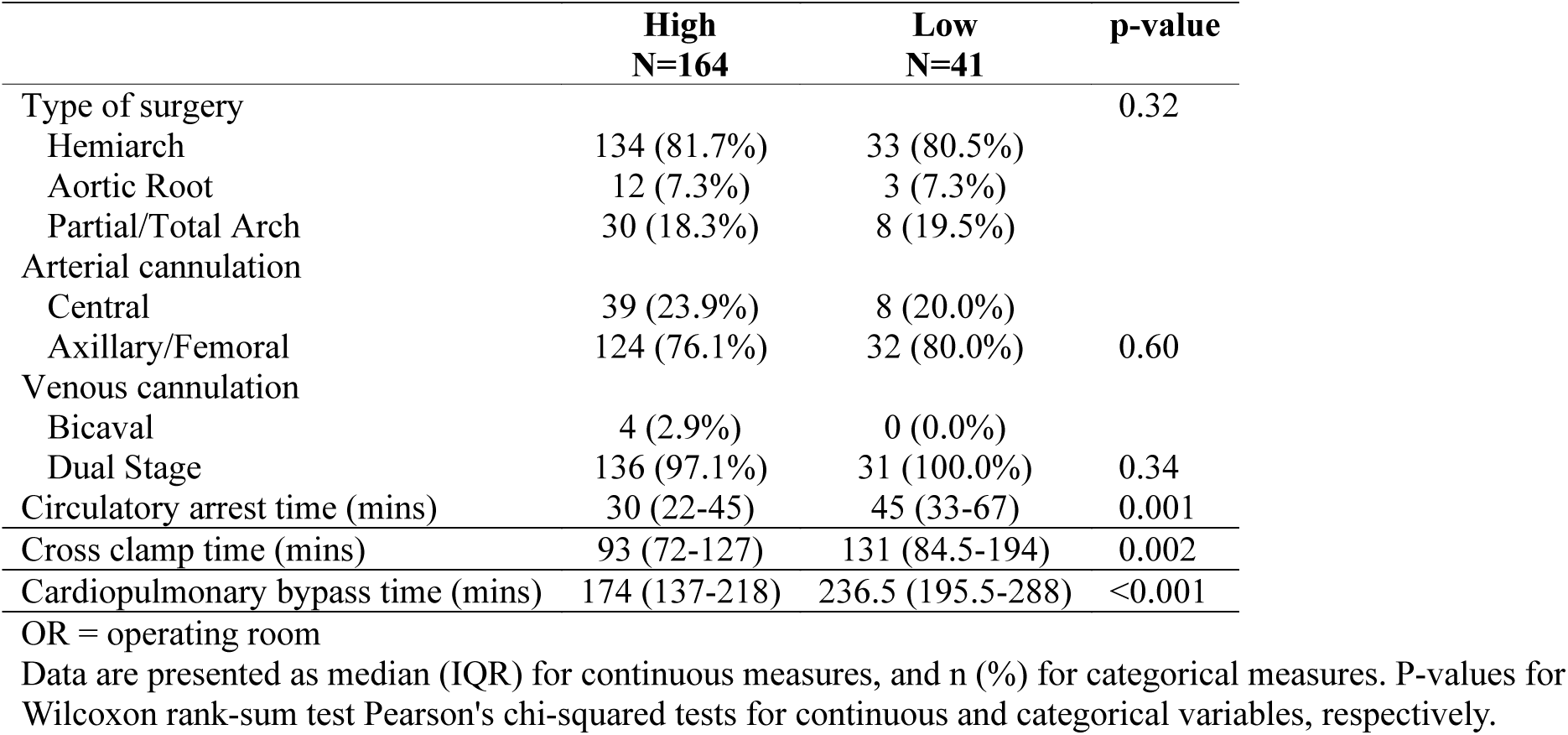
Operative Characteristics Stratified by Center Volume.

There were no statistically significant differences in the types of surgery performed. Intraoperatively, a Hemiarch was the predominant repair performed at both high and low volume centers. Between the high and low volume centers, there were an equal distribution of aortic root replacements (Modified Bentall procedure with valved conduits) performed as well as a partial (anastomosis at zone 1 or 2) and total arches. Similarly, there was not a statistically significant differences in cannulation strategy between centers (central: 23.9% vs 20.0%, p=0.60). There were only 4 [2.9%] patients at the high-volume center that required bi-caval venous cannulation to facilitate a concomitant procedure. ATAAD procedures at high-volume center were more likely to have shorter cardiopulmonary bypass (CPB) [174 (137-218) versus 236.5 (195.5-288) minutes, p <0.001], cross clamp [93 (72-127) versus 131 (94.5-194) minutes, p=0.002], and circulatory arrest times [30 (22-45) versus 45 (33-67) minutes, p=0.001] (Figure 1).

Amongst postoperative complications (Table 3), low volume centers had a higher rate of renal [12 [7.3%] versus 7 [17.1%], p=0.031] and respiratory failure [13 [7.9%] versus 8 [19.5%], p=0.029], which was statistically significant. Development of right ventricular dysfunction occurred at a statistically higher rate at the high-volume center [28 [14.1%] versus 2 [4.9%], p=0.023]. The 30-day mortality was 9.36% versus 20.6% at the high versus low volume centers, respectively [p=0.06]. The 90-day mortality was 12.36% versus 23.24% at the high versus low volume centers, respectively[p=0.07]. The univariable, survival analysis using Kaplan Meier method did show a statistically significant difference in survival at 3 years – 81.5% versus 66.7% [p=0.009] (Figure 2). Utilizing a multivariable Cox regression model, having surgery at a high-volume center was found to be associated with a statistically significant difference in 3-year survival (Adjusted Hazard Ratio (AHR): 1.21,95% Confidence Interval (CI): 1.03-1.42, p=0.021). Other factors that were found to be associated with a statistically significant difference in 3-year survival decrement included presence of coronary malperfusion (AHR: 1.96, 95% CI: 1.01-3.91, p=0.03) on presentation and cardiopulmonary bypass time (AHR: 1.02, 95% CI: 1.01-1.03, p&lt;0.001) (Table 4).

**TABLE 3.**
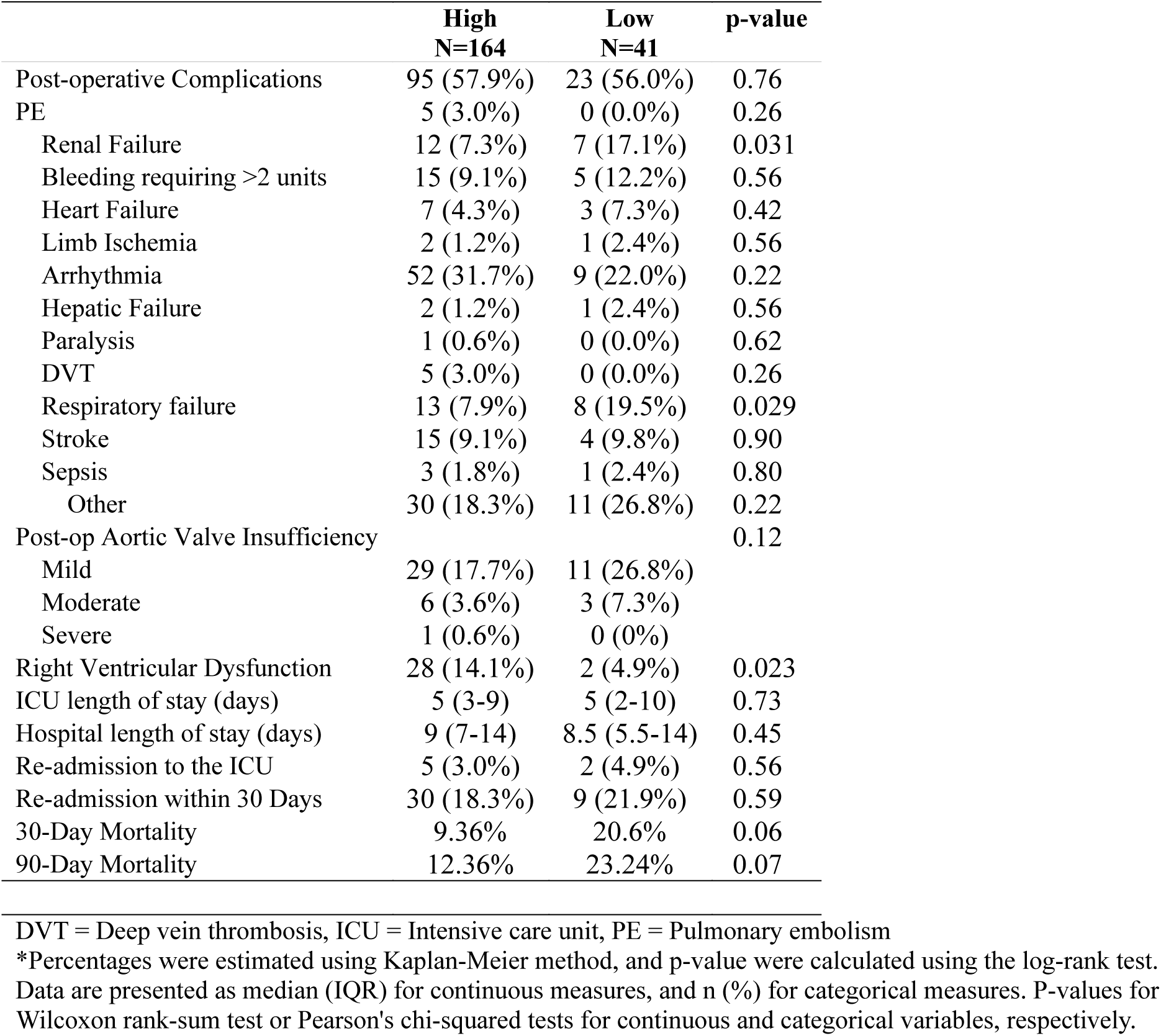
Postoperative Outcomes Stratified by Center Volume.

|  | **High**<br>**N=164** | **Low**<br>**N=41** | **p-value** |
| --- | --- | --- | --- |
| Post-operative Complications | 95 (57.9%) | 23 (56.0%) | 0.76 |
| PE | 5 (3.0%) | 0 (0.0%) | 0.26 |
| Renal Failure | 12 (7.3%) | 7 (17.1%) | 0.031 |
| Bleeding requiring >2 units | 15 (9.1%) | 5 (12.2%) | 0.56 |
| Heart Failure | 7 (4.3%) | 3 (7.3%) | 0.42 |
| Limb Ischemia | 2 (1.2%) | 1 (2.4%) | 0.56 |
| Arrhythmia | 52 (31.7%) | 9 (22.0%) | 0.22 |
| Hepatic Failure | 2 (1.2%) | 1 (2.4%) | 0.56 |
| Paralysis | 1 (0.6%) | 0 (0.0%) | 0.62 |
| DVT | 5 (3.0%) | 0 (0.0%) | 0.26 |
| Respiratory failure | 13 (7.9%) | 8 (19.5%) | 0.029 |
| Stroke | 15 (9.1%) | 4 (9.8%) | 0.90 |
| Sepsis | 3 (1.8%) | 1 (2.4%) | 0.80 |
| Other | 30 (18.3%) | 11 (26.8%) | 0.22 |
| Post-op Aortic Valve Insufficiency |  |  | 0.12 |
| Mild | 29 (17.7%) | 11 (26.8%) |  |
| Moderate | 6 (3.6%) | 3 (7.3%) |  |
| Severe | 1 (0.6%) | 0 (0%) |  |
| Right Ventricular Dysfunction | 28 (14.1%) | 2 (4.9%) | 0.023 |
| ICU length of stay (days) | 5 (3-9) | 5 (2-10) | 0.73 |
| Hospital length of stay (days) | 9 (7-14) | 8.5 (5.5-14) | 0.45 |
| Re-admission to the ICU | 5 (3.0%) | 2 (4.9%) | 0.56 |
| Re-admission within 30 Days | 30 (18.3%) | 9 (21.9%) | 0.59 |
| 30-Day Mortality | 9.36% | 20.6% | 0.06 |
| 90-Day Mortality | 12.36% | 23.24% | 0.07 |

DVT = Deep vein thrombosis, ICU = Intensive care unit, PE = Pulmonary embolism
\*Percentages were estimated using Kaplan-Meier method, and p-value were calculated using the log-rank test.
Data are presented as median (IQR) for continuous measures, and n (%) for categorical measures. P-values for Wilcoxon rank-sum test or Pearson's chi-squared tests for continuous and categorical variables, respectively.

**Table 4.** Multivariable Cox Regression Model with 3-Year Survival Following Operation.

|  | Adjusted Hazard<br>Ratio | 95% Confidence Interval | p-value |
| --- | --- | --- | --- |
| Center Volume |  |  |  |
| High | Reference | Reference | Reference |
| Low | 1.21 | [1.03-1.42] | 0.021 |
| Female | 1.45 | [0.67-3.13] | 0.34 |
| Previous Cardiac Surgery | 0.99 | [0.38-2.54] | 0.98 |
| Known Aortic Aneurysm | 1.96 | [1.01-3.91] | 0.03 |
| Lactate Level (per 1 mmol/L) | 1.14 | [0.94-1.38] | 0.15 |
| Coronary Malperfusion | 5.82 | [1.78-19.02] | <0.001 |
| Cardiopulmonary bypass time<br>(per min) | 1.02 | [1.01-1.03] | <0.001 |

## Discussion

The recent consensus from AATS, AHA/ACA, and others is to consider transfer of patients presenting with ATAAD to the nearest CAC. What’s surprising is the case volume that is needed to be considered a high-volume aortic center – some quoting only greater than or equal to 7 cases a year.^4–6^ With respect to ATAAD repair, there are many factors that determine a patient’s outcomes – preoperative characteristics (i.e. comorbidities, age, or presence of malperfusion), technical factors (e.g. cannulation strategy, CPB/circulatory arrest time, cerebral perfusion strategy, or extent of repair), and postoperative care.^10, 11^ Although the preoperative characteristics in our study were not significantly different between the two centers there were a greater percent of patients that presented with inotropic and/or pressor support [14.2% versus 4.9%], mechanically intubated [8.6% versus 2.4%], or required cardiopulmonary resuscitation [3% versus 0%] on presentation. A potential reason for this could be that our high-volume center is a referral center for hospitals in the region therefore are more likely to have patients transferred to them, due to the severity of their dissection. Despite a greater number of patients presenting in extremis (i.e. having required CPR or pressor support) at the high-volume center, this did not seem to affect the survival curves.

The sequential organ failure assessment (SOFA) score is validated to predict morbidity and mortality after cardiac surgery and has been shown to increase with higher CPB times. Even more specifically, postoperative acute renal failure has been shown to be associated with increased CPB times.^11, 12^ Surgical times can depend on the severity of the dissection or the complexity of ATAAD repair. Surgeon experience with aortic procedures can also alter how expeditious a repair can be done, which translates to changes in CPB time, clamp-time, and circulatory arrest time. There was a clinical and statistically significant difference in CPB time, cross-clamp time, and circulatory arrest time, at the high versus low volume centers. This can be due to a myriad of reasons but given there was not a statistically significant differences in type of surgery performed, with a Hemiarch being the predominant surgery along with an equal percent of root replacements being performed, we hypothesize that this may be a product of surgeon experience with aortic work. With elevated surgical times, in particular CPB and circulatory arrest times, our study showed that the low volume centers had a significantly greater postoperative renal failure and respiratory failure rate. This did not necessarily translate into longer ICU or hospital length of stays.

Aortic operative volume has been strongly associated with outcomes for ATAAD repair, being less dependent on the type of procedure performed but the overall care provided by a multidisciplinary team accustomed to treating aortic disease. Many centers across the country and part of Europe have already implemented transfer protocols to expeditiously mobilize patients to high volume centers.^13, 14^ The difference in 30- and 90-day mortality, although not statistically significant, appeared clinically impactful. The 30- and 90-day mortality was half of what it was at our low volume centers and this trend continued at 3-years, as shown with the univariable - survival analysis using Kaplan Meier method and the multivariable Cox regression model.

## Conclusion

ATAAD repair is a technically challenging procedure, but outcomes are not only dependent on surgical technique. The ability to provide comprehensive multidisciplinary care is very, if not as important, in improving survival. The results from our study mirrored published findings – which showed improved outcomes when surgery was performed by surgeons with greater experience at comprehensive aortic centers. Time from presentation to surgery influences mortality, but overall mortality has been shown to be much more dependent on where the repair is performed. A single year snapshot (2024) was able to show a mortality rate of 10% at our high-volume center, well below the national average and certainly well below our low-volume hospitals. Given this trend, further data may reveal a mort statistically relevant trend. Whenever patients present to hospitals that are not experienced aortic centers, even in the face of potential transport delays, transfer of patients to a high-volume aortic center may serve to further decrease the acute mortality and complications associated with ATAAD. In-network coordination and relationships with surrounding hospital systems are key to this process.

## Declarations

Nothing to declare.

## IRB approval

The Institutional Review Board (IRB) at Allegheny Health Network was obtained for this retrospective observational study.

## Authors’ contributions

All authors contributed equally to the generation of this article.

## Data Availability

We obtained Institutional Review Board approval for this study and all relevant data is presented in the article.

## Acknowledgements

No acknowledgements.

## Glossary

ATAAD: Acute type A aortic dissections
CAC: Comprehensive aortic center
GERAADA: German registry for acute aortic dissection type A score
ICU: Intensive care unit
SOFA: Sequential organ failure assessment

